# Cooccurrence of N501Y, P681R and other key mutations in SARS-CoV-2 Spike

**DOI:** 10.1101/2021.12.25.21268404

**Authors:** Carol Lee, Shruthi Mangalaganesh, Laurence O.W. Wilson, Michael J. Kuiper, Trevor W. Drew, Seshadri S. Vasan

## Abstract

Analysis of circa 4.2 million severe acute respiratory syndrome coronavirus 2 (SARS-CoV-2) genome sequences on ‘Global Initiative on Sharing All Influenza Data (GISAID)’ shows the spike mutations ‘N501Y’ (common to Alpha, Beta, Gamma, Omicron variants) and ‘P681R’ (central to Delta variant’s spread) have cooccurred 3,678 times between 17 October 2020 and 1 November 2021. In contrast, the N501Y+P681H combination is present in Alpha and Omicron variants and circa 1.1 million entries. Two-thirds of the 3,678 cooccurrences were in France, Turkey or US (East Coast), and the rest across 62 other countries. 55.5% and 4.6% of the cooccurrences were Alpha’s Q.4 and Gamma’s P.1.8 sub-lineages acquiring P681R; 10.7% and 3.8% were Delta’s B.1.617.2 lineage and AY.33 sub-lineage acquiring N501Y; remaining 10.2% were in other variants. Despite the selective advantages individually conferred by N501Y and P681R, the N501Y+P681R combination counterintuitively didn’t outcompete other variants in every instance. Although a relief to worldwide public health efforts, in vitro and in vivo studies are urgently required in the absence of a strong in silico explanation for this phenomenon. This study demonstrates a pipeline to analyse combinations of key mutations from public domain information in a systematic manner and provide early warnings of spread.

## 1. Introduction

The severe acute respiratory syndrome coronavirus 2 (**SARS-CoV-2**), which causes the ongoing novel coronavirus disease 19 (**COVID-19**) pandemic, has a single-stranded positive-strand ribonucleic acid genome (ssRNA(+) genome) of size 26–32 kb, with high fidelity replication due to 3′-to-5′ exoribonuclease ‘proof-reading’ mechanism [1,2]. As this virus adapts to its human host, we have seen it evolve and present quasispecies diversity [2,3]. While most of the several thousands of mutations catalogued to date aren’t substantial functional changes they have proven aetiologically useful [4,5].

Two years on since the start of the COVID-19 pandemic, we now have a good idea of the key mutations, especially in the Spike protein, which are punctuations in the evolutionary story of this virus to date. The D614G mutation reported in April 2020 resulted in the ‘G-strain’ with increased infectivity replacing the genomic background of this virus globally, although there was no impact on vaccine efficacy [5,6,7]. However, the N501Y mutation common to the Alpha, Beta, Gamma and Omicron variants of concern (**VOC**), has contributed to enhanced infection and transmission, reduced vaccine efficacy, and the ability of SARS-CoV-2 to infect new species such as wild type mice [8,9,10,11]. Another key mutation is the P681R which alters the furin cleavage site, and has been responsible for increased infectivity, transmission and global impact of the Delta variant [12,13,14].

Our primary objective is to investigate the risk of the three aforementioned ‘mutations of current interest’ (**MOCI**) cooccurring naturally due to convergent evolution and resulting in a SARS-CoV-2 variant that is of greater concern than those declared to date, noting that the latest Omicron VOC has the N501Y but not the P681R mutation. Our secondary objective is to demonstrate a methodology and pipeline to analyse the spread of variants containing combinations of important mutations. We have achieved this by mining data from **GISAID**, the Global Initiative on Sharing All Influenza Data, which is the largest and the most comprehensive repository of SARS-CoV-2 sequences [15,16]. We have used quasispecies theory and in silico modelling to interpret our findings to the extent possible, and recommended future research directions.

## 2. Results: Frequency and distribution of D614G, N501Y and P681R mutations

The earliest record of these three MOCI coming together was in Slovenia on 17 October 2020, however, no further observations were recorded since then in that country. Mining of GISAID data found 3,678 entries (0.1%) containing the MOCI and a majority of these were in current VOC – Alpha (61.3%), Beta (0.4%), Gamma (6.4%), Delta (21.2%), Omicron (0.0%) – and a small proportion in current or former VOI – Mu (0.3%), Kappa (0.2%) and Iota (0.02%). A small proportion (10.2%) was also observed in other variants not classified as VOC, VOI, Variant Under Monitoring (VUM), or specific sub-lineages (**Figure 1**).

**Figure 1.**
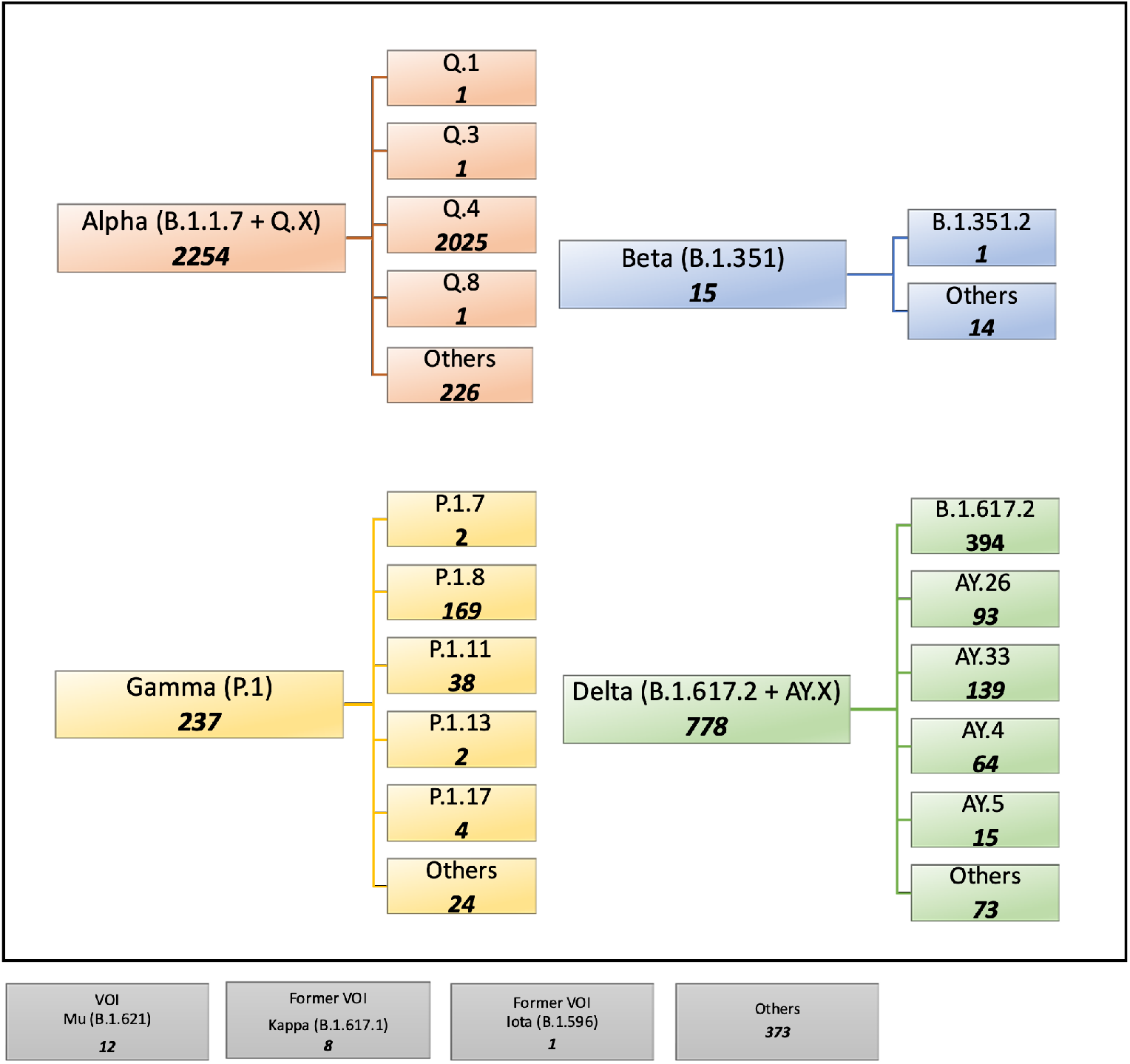
Summary of SARS-CoV-2 lineages and sub-lineages containing the D614G, N501Y and P681R mutations of current interest (MOCI) on GISAID. 3,678 sequences were found with our MOCI from a total of 4,177,098. Bold numbers in each box shows the number of observations for each VOC/VOI and their sub-lineages (where such information was available). ‘Others’ indicate samples not further classified on GISAID.

**Figure 2** illustrates the MOCI frequency in countries with at least 50 recorded sequences – Brazil (5.8%), Denmark (1.9%), France (22.7%), Germany (3.6%), Sweden (5.2%), Turkey (21.4%), UK (3.6%) and USA (22.1%). These eight countries represent 86.4% of the cases containing the MOCI (**Figure 3A**, which shows a continuous timeline). Fifty-seven other countries recorded less than 50 entries for the period 17 October 2020 to 15 September 2021 (**Supplementary Table S1**).

**Figure 2.**
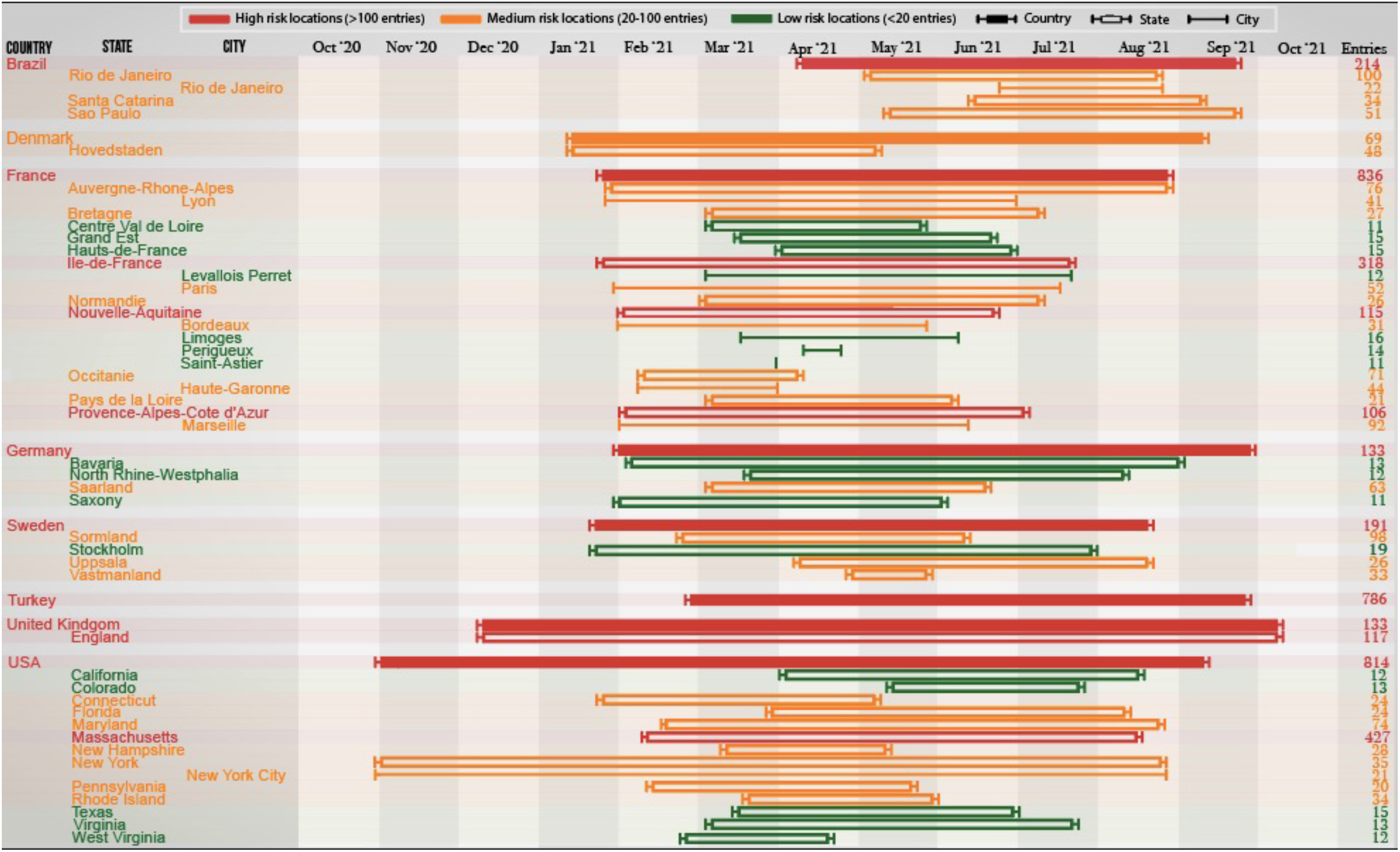
Discrete timeline depicting the start and end which SARS-CoV-2 variants with the P681R, N501Y and D614G mutations were observed in various countries, states and cities between October 2020 and October 2021. Countries with 50 or more observations, and states/cities with 10 or more observations are deemed significant and thus included in this timeline. Raw data with the full list of countries are provided in **Supplementary Table S1**.

Three countries – France, USA and Turkey – stand out as they each contribute to over 22% of the total instances of the MOCI cooccurring. It’s unsurprising that these trends overlap with the spread of VOC in these nations (**Figures 3B-3D**), because there would have been more opportunities for Delta to acquire N501Y, and for Alpha or Gamma to acquire P681R (**Figures 3B-3F**). Such cooccurrences also appear to have happened over short periods of time, for instance during March to May 2021 in France and USA with the Alpha VOC; and between June and August 2021 in Turkey with the Delta VOC (**Figures 2 and 3B-3D**). This could be due to founder effects, but we cannot establish this by mining GISAID data alone; it will require detailed epidemiological investigations by national public health authorities, which is beyond the scope of this work. Nevertheless, from **Figure 2,** we can be reasonably sure that a number of independent events of convergent evolution (i.e. MOCI cooccurring) have taken place. It is worth investigating why we do not see as many instances of Delta acquiring N501Y in France and USA, even though they had a very high number of this VOC from July 2021.

**Figure 3.**
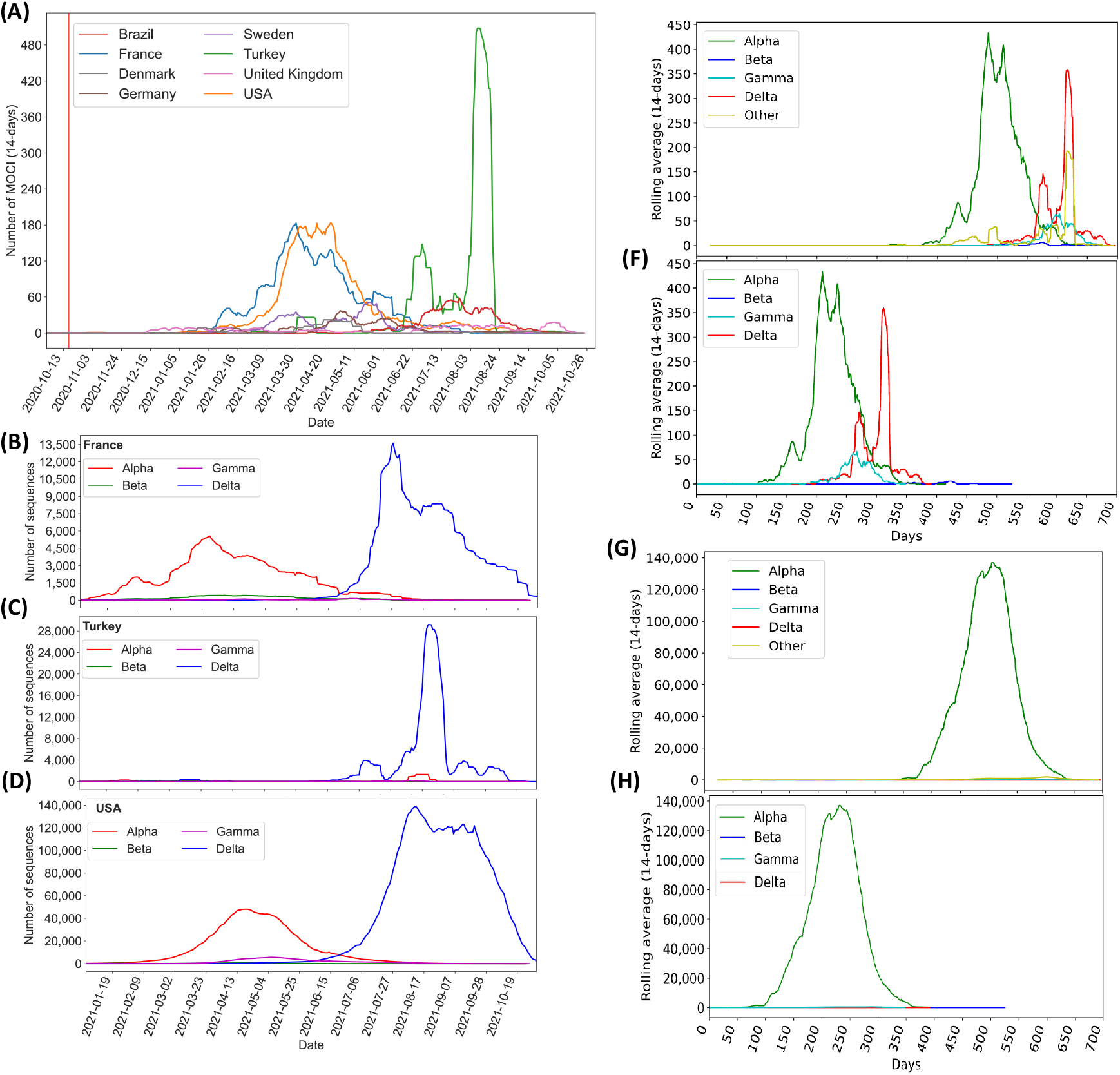
Continuous representation of the timeline during which SARS-CoV-2 variants with the P681R/H, N501Y and D614G mutations were observed between October 2020 and October 2021. **(A)** Number of isolates with MOCI, plotted as 14-day rolling average, in the eight countries which had at least 50 observations. The vertical red line shows the first recorded cooccurrence in Slovenia on 17 October 2020; **(B-D)** Number of VOC sequences observed in the top three countries (France, Turkey, USA respectively); **(E-F)** Number of isolates with MOCI which are also VOC, plotted as 14-day rolling average, either from the notional start of the pandemic in December 2019, or from September, May, November and October 2020 as the first reported months for Alpha, Beta, Gamma and Delta respectively. **(G-H)** Number of isolates with the P681H (rather than P681R), N501Y and D614G, plotted as 14-day rolling average, either from the notional start of the pandemic in December 2019, or from the month of first report for each VOC. ‘Other’ indicates MOCI found in variants other than these VOC.

## 3. Discussion

### 3.1. Y501-R681 almost always in the G614 background

From **Supplementary Table S2** we see that 3,688 entries contain N501Y+P681R, just ten more than the 3,678 entries that contain D614G+N501Y+P681R; in other words, the instances of N501Y cooccurring with P681R have almost always happened on the D614G background as predicted [5,6,7]. Although these three mutations are positioned sufficiently far apart in the 3-dimensional protein structure to suggest they don’t interact (**Figure 4**), further studies are required to understand whether there may be indirect functional links that could enhance viral efficiency.

**Figure 4.**
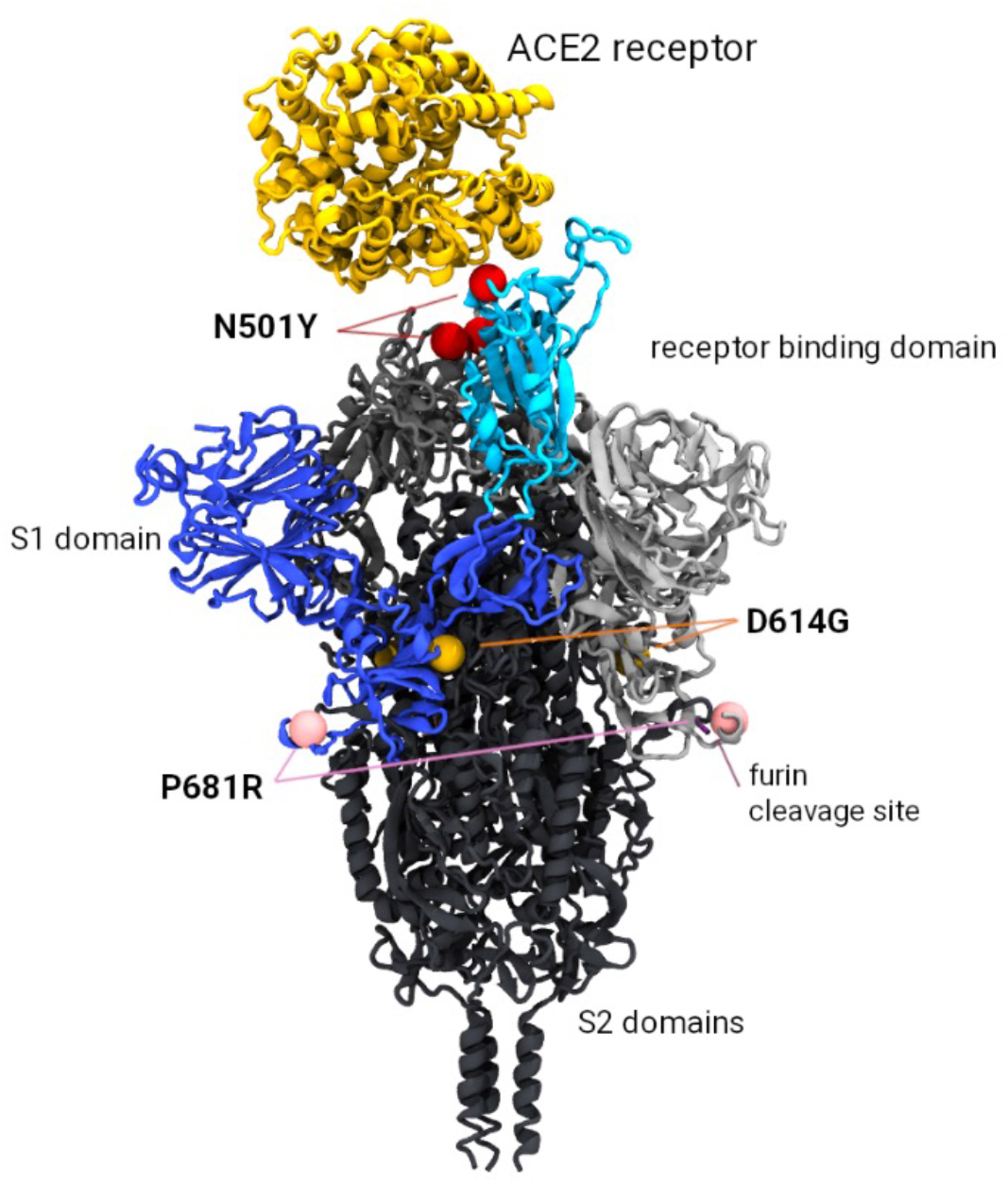
Model of the SARS-CoV-2 Spike trimer protein indicating the relative position of the P681R, N501Y and D614G mutations of current interest (MOCI). The N501Y mutation occurs in the receptor binding domain (highlighted in cyan) and is associated with increased binding affinity to the ACE2 receptor (shown in yellow). The P681R mutation is implicated with more efficient cleavage of the S1/S2 furin site (required prior to viral fusion to the host cell). Mutation D614G occurs at the S1/S2 interface and has also been implicated with increased replication efficiency.

### 3.2. Predominance of Alpha Q.4 acquiring P681R and Delta acquiring N501Y

We see that the cooccurrence of the MOCI is largely due to; (a) the Alpha VOC acquiring the P681R (61.3%), and this has happened overwhelmingly in Alpha’s Q.4 sub-lineage (89.9%) by definition; (b) the Delta VOC acquiring the N501Y (21.2%), and this has mainly happened in Delta’s parent lineage (B.1.617.2; 50.6%) and also the sub-lineage AY.33 (17.9%); (c) the Gamma VOC acquiring the P681R (6.4%), predominantly in its P.1.8 sub-lineage (71.3%). It is worth noting that the Q.4 sub-lineage has the P681R by definition (https://outbreak.info/situation-reports?pango=Q.4). The Beta VOC acquiring the P681R only constituted 0.4% of the total occurrences, and this was almost entirely in the B.351.2 parent lineage (93.3%). **Supplementary Table S3** shows the results for other key Spike mutations in comparison to those obtained from GISAID. For H69del and Y145del, more samples were identified using our pipeline than on GISAID – 1 138 302 versus 1 123 439 before date filtering and 2 563 versus 1 364 after date filtering

### 3.3. Alpha acquiring P681R ahead of other VOC could be due to P/H681R

Alpha, Beta and Gamma each have N501Y, however Alpha was the first VOC to acquire P681R, both in terms of absolute (**Figure 3E**) and relative (**Figure 3F**) timelines, although Beta was reported four months before Alpha in May 2020. Alpha, especially its Q.4 sub-lineage, also contributes to most instances of the MOCI cooccurring. There is a possible biomolecular basis for this; P681H is present in Alpha, but not in the other two VOC. The Grantham scores associated with P681R (103) and P681H (77) are comparable and the H681R substitution is thus a conservative change (Grantham distance 29). There is only one way to get to Histidine (H) or Arginine (R) from Proline (P); and for H and R there is only one way to get to each other; and we see no structural reason from our in silico model as to why one should be preferred to the other. Thus, we could infer that some of the instances of Alpha acquiring P681R could have been due to a substitution of H with R, which is the signature of the Q.4 sub-lineage. Spike’s 681st position is the fifth substrate sequence for cleavage recognition; both furin and the transmembrane serine protease 2 (TMPRSS2) cleave the Spike at 685/686 position with H (and possibly R) enhancing this process [7,17,18]. It is worth noting that to penetrate host cells, the SARS-CoV-2 Spike protein must be cut twice by host proteins. In the SARS-CoV-1 (SARS), both incisions occur after the virus has locked on to a cell. But with SARS-CoV-2, the presence of the furin cleavage site enables host enzymes like furin to make the first cut as newly formed viral particles emerge from an infected cell. These pre-activated viral particles can then go on to infect cells more efficiently compared to particles requiring two cuts. Thus, the P681R increases the susceptibility of the furin cleavage site in Delta VOC, and allows the exposure of the Spike’s S2 subunit for better cell integration.

### 3.4. Cooccurrences have been reported predominantly in eight countries

Cooccurrences of the MOCI were observed with Alpha, Beta, Gamma and Delta VOC in 41, 8, 11 and 47 countries respectively, indicating convergent evolution, especially as much of the world was under lockdown during 2020-2021. Curiously, two thirds of the observations have been reported from just France, Turkey and USA; 86.3% from these three countries plus Brazil, Denmark, Sweden, UK and Germany (**Supplementary Table S1; Supplementary Figure S4**). Several instances involve proximal regions, suggesting multiple founder effects (**Figures 2 and 5**). For example, in Denmark and Sweden, variants with the MOCI were concentrated in highly populous Hovedstaden and Svealand regions (Sörmland, Stockholm, Uppsala and Västmandland) [19]; the MOCI frequencies were also correlated with the most common lineage in respective countries and regions, suggesting community transmission, although these MOCI did not go on to become the dominant variant in those locations. This is in line with most within-host variants getting lost during transmission, and only a few founding infections being maintained in a given population [20].

**Figure 5.**
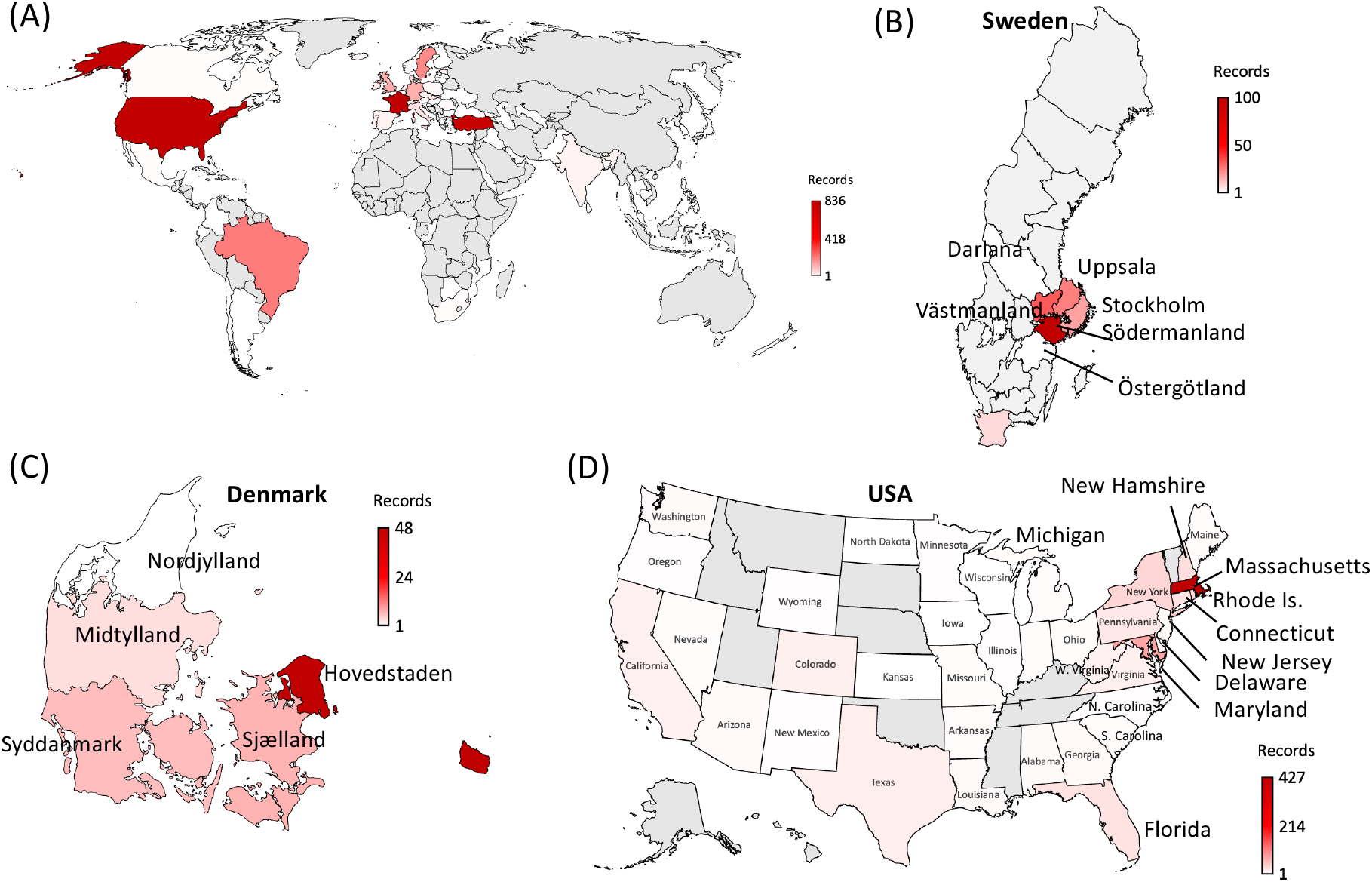
World map depicting the 65 countries where SARS-CoV-2 variants containing the cooccurring MOCI were recorded between October 2020 and October 2021. **(A)** world map, with zoomed in view of the regions observed with MOCI shown for **(B)** Sweden **(C)** Denmark and **(D)** USA. Countries are color-coded based on the number of records of MOCI observed, as shown in the legend, with greyed out regions indicating no observations. For some countries such as Turkey, region/city-level information was not available, but EP _ISL numbers can be provided upon request.

Cooccurrence of the MOCI in the UK occurred mostly with the Alpha VOC’s B.1.1.7 and Q.4 from late 2020 to May 2021; followed by the second wave of the Delta VOC since April 2021, especially in AY.4 that had the highest (58%) prevalence (**Figure 2**). The former period had ten times the death rate (∼1200 per week between September 2020 and March 2021) compared to the latter (∼120 per week; probably due to lockdowns and 30% of the population already double-vaccinated). In the UK which has a very high rate of sequencing, the overall numbers of cooccurring MOCI were still low (133 records, 88% in England). Of these, there were respectively 52, 39 and 18 instances of the AY.4 acquiring N501Y, and the B.1.1.7 and Q.4 acquiring P681R [21,22]. It is possible that the lockdown protocols and vaccination coverage may have influenced the observed viral transmission dynamics [23]. In the Americas, most of 1,078 cooccurrences of the MOCI were detected either in the US East Coast (673 entries), or in the Sul (Santa Catarina) and Sudeste (São Paulo, Rio de Janeiro) regions in the South and Southeast of Brazil (284 entries). These trends could be due to founder effects and lack of travel restrictions from early 2021, especially in the United States [24] (**Figures 2 and 5**). In Brazil, the cooccurrences of the MOCI also corresponded with the prevalence of P.1 until August 2021 after which the AY.99.2 became the dominant variant (**Supplementary Figure S4**). Unfortunately, regional and city level information was not available for Turkey on GISAID, preventing further analysis and insights and highlighting the importance of metadata [16].

### 3.5. Y501-R681 doesn’t outcompete other variants

From our analysis, SARS-CoV-2 isolates containing Y501-R681 in Spike did not outcompete other variants in every single instance we have examined (**Figures 2 and 3**), which is both counterintuitive and a relief to worldwide public health efforts. Although individually these two mutations have been reported to confer selective advantage [8,9,10,11,12,13,14], their combination apparently has not and we have not found any compelling biomolecular reason for this (**Figure 4**). It would be desirable to conduct in vitro and in vivo studies to gain a better understanding, either using infectious clones and reverse genetics [25,26,27,28], or using naturally occurring comparable isolates with and without these mutations [see for instance 7]

### 3.6. Implications for the Omicron VOC

Alpha and Omicron each have the Y501-H681 combination which is present in 1,085,434 entries out of the 4.2 million we have examined in this study (i.e. over 25%); this contrasts with the Y501-R681 of which we have only found 3,688 observations. Just as the Q.4 sub-lineage with Y501-R681emerged when the Alpha VOC started to spread, it is possible that a new sub-lineage of Omicron with Y501-R681 could emerge given the rapid rise of this latest VOC [29,30,31]. In this context, it is important to highlight the problem of sequencing artefact, for example 65 high coverage sequences of Omicron with P681R recently appeared on GISAID but were subsequently corrected by the submitting laboratory following repeat libraries with fewer cycles and other improvements [32]. The spread of VOC in immunocompromised patients could lead to mutations with a selective advantage for antibody escape and/or transmissibility (e.g. N501Y and P681H/R), in addition to a number of deletions in the N terminal domain [33], and this aspect needs further investigation given Omicron’s large number of mutations (many of them yet to be studied in depth). The adaptation and evolution of this virus in new hosts, for instance mice and rats, also pose additional risks such as additional reservoirs and reinfection of humans [11]. Investigations into the dynamics affecting the SARS-CoV-2 pandemic in each of the eight countries are also warranted, in the wider context of travel restrictions, population demographics and dynamics [34,35]. Lack of patient-deidentified metadata in a consistent format further complicates meaningful analysis as emphasized before [16].

### 3.7. SARS-CoV-2 evolution and selective pressures on quasispecies

Any virus which is new to a host will undergo a period of adaptation, as numerous selective pressures come to bear. RNA viruses, such as coronaviruses, are error-prone in their replication and exist as a cloud of variants, with the dominant clade representing the majority population, but with a number of diverse variants also represented within the population, known as quasispecies [2,3,36,37]. Among viruses which are established in their host, the dominant clade is generally relatively ‘fit’ in its ability to replicate and out-competes all the other clades. However, when a virus is new to a host, its relative fitness may be quite low, so, in any particular environment, different clades can have similar relative fitness. This means subtle differences might give a particular clade a small advantage enabling it to predominate as the population continues its host adaptation. The environment may comprise of host factors, such as host genetics, tissue tropism, age, immune status or competence, presence of other infections, as well as external factors, such as temperature and humidity. With SARS-CoV-2, the extraordinary public health measures in some regions of the world are likely exerting their own selective pressures on its evolution and creating numerous local environments. This phenomenon, called the ‘survival of the flattest’ [3] can lead to the emergence of multiple clades dominating under different environments. These is also an increasing appreciation that, within the host, different clades may predominate in different tissues at the same time and that there may be interactions among the diverse quasispecies that facilitate the infection [38].

The pathway of adaptation is not always towards severe disease, since this may not lead to maximum replication and transmission. Indeed, many viruses with a long history of acute infection in an established host species, the disease may be mild or inapparent, reflecting a relationship more towards stasis, where the impact on the host does not compromise the chances of the progeny virus being transmitted to a new host. For many coronaviruses, we have seen an evolution towards milder infection, albeit with greater transmission rates than has been seen with SARS-CoV-2 to date. Certainly, some level of reduction in the efficacy of vaccine-induced antibody in neutralising the Delta and Omicron variants has been demonstrated, but the other feature of these viruses, which is less often discussed, is the ability of these VOC to replicate by means of syncytial formation, rather than the simple apoptotic cycle seen with earlier variants [39]. This may confer an important immune evasive aspect to replication, which might offer a distinct advantage, in immune avoidance. It might also explain why these clades are more productive and less pathogenic since systemic infection is not always involved.

### 3.8. Error catastrophe, Muller’s ratchet and implications for SARS-CoV-2

A phenomenon related to quasispecies evolution is that of error catastrophe, which may occur if the mutational rate exceeds the ability of the fitness landscape to accommodate the resultant evolving clades [40]. The key triggers for this are not fully understood, but it is thought that the application of strict biosecurity measures may play a part by restricting the availability of new hosts. In such cases, the quasispecies becomes increasingly attenuated, with mutations accumulating via a mechanism called ‘Muller’s ratchet’, to the point where the epidemic may die out [41]. This has been seen in many prior outbreaks of Ebola, where the infection became increasingly less pathogenic and less transmissible. While the current environment of SARS-CoV-2 is complex and highly variable, the selective pressure on a virus is towards higher transmissibility and milder disease in all environments, and the current systems of control preferentially select for such. Once a virus develops either/both of these traits, it is very unusual for the reverse to occur because viruses with higher transmissibility out-compete those with lesser transmissibility and those which are milder disable the host less, so they continue to travel and mix with susceptible hosts. It remains to be seen if this is being observed with the Omicron VOC as early reports indicate higher transmissibility and less severe disease.

In comparing variants of SARS-CoV-2, it is tempting to focus on specific mutations and their location – also perhaps with overt attention being paid to the Spike protein, especially its receptor binding domain – and not consider that the observed changes could evolve independently of each other. The secondary and tertiary folding of proteins, as well as post translational modifications, can have a profound effect on function, such that a mutation at one site might require a complimentary change at another site in order to confer advantage, or be wiped out by another, seemingly unrelated change. For the same reason, convergent evolution, might also occur, exhibited by the co-existence of identical co-mutations in virus clades of different lineage [42]. Another phenomenon known to occur in many coronaviruses is recombination, where partial genomic exchange may occur between two coronaviruses when simultaneously infecting the same host. The extent to which this may have occurred in SARS-CoV-2 is the likely subject of future analysis [43,44].

## 4. Methods

### 4.1. Bioinformatics analysis

We mined GISAID data on 3 November 2021 containing 4,835,087 entries, 99.9% (4,831,865) sequenced from people who tested COVID-19 positive. Of these, 4,748,462 contained over 29 thousand nucleotides and were deemed ‘complete’. GISAID has also defined a subset of these 4.8 million sequences, containing 3,519,085 entries, as ‘high coverage’. This is because 99% of their bases are defined; there is no unverified deletion or insertion; and unique mutations not seen in other sequences constitute fewer than 0.05% in these entries. Consequently, some of our analysis of the data downloaded on 1 November 2021 (4,766,139 sequences) required manual curation whenever we encountered incorrect dates, or low coverage sequencing that could skew our results. **Supplementary Table S5** provides GISAID’s break down of these 4.8 million entries by variants containing the D614G (98.8%), N501Y (27.01%) and P681R (48.2%) mutations; by Alpha (23.9%), Beta (0.8%), Gamma (2.4%), Delta (47.8%), Omicron (0.0%) VOC; and by Lambda (0.2%) and Mu (0.3%) variants of interest (**VOI**).

Human-origin SARS-CoV-2 sequences over 29 kb length were aligned to the nominal reference (EPI_ISL_402123) using an in-house alignment pipeline that generated a file in the ‘variant call format’ (**VCF**) containing all the mutations as of 1 November 2021 [1,2]. In the VCF containing 4,294,469 genome sequences, we searched for entries with MOCI, for example, the Spike N501Y was searched at nucleotide position 23403 for an A to G transition. The EPI_ISL sample IDs containing our MOCI were merged with their respective GISAID metadata (location, date of collection, lineage) to create an annotated database. **Supplementary Table S5** shows the split up of these 4.3 million sequences by variants containing the D614G (98.8%), N501Y (29.2%) and P681R (44.2%) mutations; and by Alpha (26.3%), Beta (0.9%), Gamma (2.2%), Delta (43.8%), Omicron (0.0%), Lambda (0.2%) and Mu (0.3%) VOC/VOI.

Using a custom Python code, we identified the presence of the D614G, N501Y and P681R mutations, and combinations thereof (c.f. ‘Data Availability Statement’ for our code). Our final dataset contained 4,177,098 entries after filtering out GISAID records with incomplete dates, i.e. retaining those with a ‘YYYY-MM-DD’ format. We ascertained whether these isolates are one of the VOC (and if yes, which sub-lineage), their location (country, state, and city depending on information available), and sample collection date. Using the latter, we calculated rolling averages over 14-days, as this window has been shown from previous experience to be an optimal period to reduce background noise, especially as the data is discrete and highly variable in size across countries [11]. However, unlike our previous study in which we were interested in macroscopic trends, here we have looked at every instance of key mutations cooccurring; therefore, we did not use a threshold or filter based on minimal sample size. Our analysis presents the spread of these mutations from the notional start of the SARS-CoV-2 pandemic (31 December 2019) to the end of our observation period (01 November 2021) with appropriate classifications (e.g. Alpha, Beta, Gamma, Delta).

### 4.2. Biomolecular modelling

Fully glycosylated in silico models of the SARS-CoV-2 Spike protein were constructed based on the ‘6VSB’ protein databank (PDB) structure [45], minus the transmembrane domain (residues 1161-1272) for additional computational efficiency as described previously [ 7]. Models were simulated in aqueous solution (TIP3, water, 0.15M ions, NVT ensemble, 310K) using NAMD 2.14 software [46]. Models were visualized with the visual molecular dynamics (VMD) program for large biomolecular systems which uses 3-D graphics and built-in scripting to assess the spatial arrangement of mutations and any complementary or inhibitory interactions [47].

## 5. Conclusions and limitations

SARS-CoV-2 is the most sequenced virus in the world, however, there is an inherent bias introduced by the highly variable sequencing-to-COVID-19 positivity ratio (**S:P**) across countries (e.g., the UK has one of the highest P:S ratios in the world), and across time (S:P has generally gone up in all countries, e.g. India). Thus, the GISAID data is likely to contain non-random samples with a skew in favour of sequences associated with epidemiologically consequent cases, albeit this is difficult to decipher as most sequences have little or no meaningful patient-deidentified information. The lockdowns and vaccination coverage associated with this viral pandemic have also been unprecedented in human history and spatio-temporally variable, further complicating our analysis. Notwithstanding these limitations, which have led to the proliferation of certain clades in certain environments, we are still able to make broad conclusions thanks to the very large number of sequences available on GISAID. We are also able to demonstrate a process and pipeline to analyse mutations of structural, functional and epidemiological consequence from public domain information in a systematic manner and provide early warnings of certain combinations starting to spread. Though it is unclear which combinations of mutations provide the best selective advantage, there is concern that mutations previously observed in other dominant lineages may arise spontaneously in contemporary strains further increasing their potential for infectivity and adverse clinical outcomes. In this case-study we analysed three key mutations (viz. D614G, N501Y and P681R) cooccurring but found no evidence of this combination spreading. Although 3,678 sequences were found on GISAID between 17 October 2020 to 1 November 2021, mainly in France, Turkey and USA, the Y501-R681 combination hasn’t outcompeted other variants and this warrants further in silico, in vitro and in vivo investigations. With the latest VOC Omicron containing a large number of mutations, many yet to be studied in-depth, our methodology will be very useful to understand whether certain combinations of mutations are more transmissible. If GISAID entries are double-checked for sequencing artefact before submission and strengthened with patient-deidentified metadata, then this approach could enable early epidemiological intelligence, for instance on case severity, mortality, and factors such as age, gender, race and co-morbidities that could increase the infection risk.

## Data Availability

This paper only uses publicly accessible data that can be downloaded from GISAID, the largest repository of SARS-CoV-2 genome sequences. The lists of GISAID EPI_ISL_ sample IDs and data used by our analysis can be requested from the corresponding author. Our Python code is available at https://github.com/Carol-Lee-gh/Covid-Mutation-Pipieline.git.

## Supplementary Materials

The following supporting information can be downloaded at: www.mdpi.com/xxx/s1, Table S1: Raw data for countries with cooccurring P681R, N501Y and D614G mutations. www.mdpi.com/xxx/s2, Table S2: Observation of key mutations and their combinations on GISAID. www.mdpi.com/xxx/s3, Table S3: Observation of other key SARS-CoV-2 Spike mutations. www.mdpi.com/xxx/s4, Figure S4: Frequency of the P681R, N501Y and D614G mutations cooccurring in the eight countries with more than 50 observations between October 2020 and October 2021. www.mdpi.com/xxx/s5, Table S5: GISAID data on key SARS-CoV-2 mutations classified by VOC/VOI.

## Author Contributions

Conceptualization and methodology, S.S.V.; bioinformatics, C.L. and L.O.W.W.; biomolecular modelling and interpretation, M.J.K. and T.W.D.; data analysis and original draft preparation, S.M., C.L. and S.S.V.; funding acquisition, S.S.V. and L.O.W.W.; manuscript review and editing, all authors.

## Institutional Review Board and Informed Consent Statements

Not applicable for the following reasons. This study does not involve any human samples, identifiable human data or patient records. It only involves bioinformatics analysis of novel coronavirus genome sequences openly accessible from the public repository ‘GISAID’, and these sequences do not contain any metadata that can be identified with any individual. This includes sample collection dates and anonymous sample IDs that are openly accessible. Our study does not report any exact age or photographs. Clinical trial registration is not applicable as this is neither an interventional study nor an observational study involving humans.

## Data Availability Statement

This paper only uses publicly accessible data that can be downloaded from GISAID, the world’s largest repository of SARS-CoV-2 genome sequences. The lists of GISAID’s EPI_ISL_ sample IDs and data used by our analysis can be requested from the corresponding author. Our Python code is available at https://github.com/Carol-Lee-gh/Covid-Mutation-Pipieline.git.

## Acknowledgments

We are grateful for support from our CSIRO colleagues at the Australian Centre for Disease Preparedness (https://www.grid.ac/institutes/grid.413322.5), especially Kim Blasdell, Simran Chahal, Alexander McAuley and Nagendrakumar Singanallur, and the Australian e-Health Research Centre, especially Denis Bauer, David Hansen, Yatish Jain, Brendan Hosking and Aidan Tay. We thank Professor Adriana Heguy of New York University Langone Health for discussions on sequences of Omicron with P681R.

---

**Supplementary Table S1.** Raw data for countries with cooccurring P681R, N501Y and D614G mutations. Frequencies for each region and continent are provided as sub-totals and totals respectively.

| Continent | Region | Country | Frequency | Sub-totals | Totals |
| --- | --- | --- | --- | --- | --- |
| Africa | Eastern Africa | Reunion | 2 | 2 | 15 |
| Africa | Sub-Saharan Africa | Botswana | 2 | 13 |  |
| Africa | Sub-Saharan Africa | Malawi | 2 |  |  |
| Africa | Sub-Saharan Africa | Mozambique | 1 |  |  |
| Africa | Sub-Saharan Africa | Republic of the Congo | 1 |  |  |
| Africa | Sub-Saharan Africa | South Africa | 7 |  |  |
| Americas | Latin America and the Caribbean | Argentina | 1 | 259 | 1078 |
| Americas | Latin America and the Caribbean | Brazil | 214 |  |  |
| Americas | Latin America and the Caribbean | Chile | 2 |  |  |
| Americas | Latin America and the Caribbean | Colombia | 1 |  |  |
| Americas | Latin America and the Caribbean | Czech Republic | 23 |  |  |
| Americas | Latin America and the Caribbean | Ecuador | 5 |  |  |
| Americas | Latin America and the Caribbean | Martinique | 4 |  |  |
| Americas | Latin America and the Caribbean | Mexico | 5 |  |  |
| Americas | Latin America and the Caribbean | Puerto Rico | 4 |  |  |
| Americas | Northern America | Canada | 5 | 819 |  |
| Americas | Northern America | USA | 814 |  |  |
| Asia | Central Asia | Uzbekistan | 1 | 1 | 856 |
| Asia | Eastern Asia | Japan | 4 | 5 |  |
| Asia | Eastern Asia | South Korea | 1 | 11 |  |
| Asia | South-eastern Asia | Cambodia | 2 |  |  |
| Asia | South-eastern Asia | Malaysia | 1 |  |  |
| Asia | South-eastern Asia | Philippines | 4 |  |  |
| Asia | South-eastern Asia | Singapore | 3 |  |  |
| Asia | South-eastern Asia | Thailand | 1 |  |  |
| Asia | Southern Asia | Bangladesh | 5 | 28 |  |
| Asia | Southern Asia | India | 21 |  |  |
| Asia | Southern Asia | Pakistan | 1 |  |  |
| Asia | Southern Asia | Sri Lanka | 1 |  |  |
| Asia | Western Asia | Georgia | 3 | 811 |  |
| Asia | Western Asia | Iraq | 1 |  |  |
| Asia | Western Asia | Israel | 18 |  |  |
| Asia | Western Asia | Turkey | 786 |  |  |
| Asia | Western Asia | United Arab Emirates | 3 |  |  |
| Europe | Eastern Europe | Bulgaria | 2 | 73 | 1728 |
| Europe | Eastern Europe | Hungary | 2 |  |  |
| Europe | Eastern Europe | Poland | 12 |  |  |
| Europe | Eastern Europe | Romania | 10 |  |  |
| Europe | Eastern Europe | Slovakia | 45 |  |  |
| Europe | Eastern Europe | Ukraine | 2 |  |  |
| Europe | Northern Europe | Denmark | 69 | 472 |  |
| Europe | Northern Europe | Estonia | 26 |  |  |
| Europe | Northern Europe | Finland | 2 |  |  |
| Europe | Northern Europe | Iceland | 8 |  |  |
| Europe | Northern Europe | Ireland | 38 |  |  |
| Europe | Northern Europe | Latvia | 1 |  |  |
| Europe | Northern Europe | Lithuania | 2 |  |  |
| Europe | Northern Europe | Norway | 2 |  |  |
| Europe | Northern Europe | Sweden | 191 |  |  |
| Europe | Northern Europe | United Kingdom | 133 |  |  |
| Europe | Southern Europe | Croatia | 12 | 117 |  |
| Europe | Southern Europe | Greece | 7 |  |  |
| Europe | Southern Europe | Italy | 47 |  |  |
| Europe | Southern Europe | Malta | 1 |  |  |
| Europe | Southern Europe | Portugal | 28 |  |  |
| Europe | Southern Europe | Slovenia | 1 |  |  |
| Europe | Southern Europe | Spain | 21 |  |  |
| Europe | Western Europe | Austria | 8 | 1066 |  |
| Europe | Western Europe | Belgium | 21 |  |  |
| Europe | Western Europe | France | 836 |  |  |
| Europe | Western Europe | Germany | 133 |  |  |
| Europe | Western Europe | Luxembourg | 8 |  |  |
| Europe | Western Europe | Netherlands | 22 |  |  |
| Europe | Western Europe | Switzerland | 38 |  |  |
| Oceania | Australia and New Zealand | New Zealand | 1 | 1 | 1 |

**Supplementary Table S2.** Observation of key mutations and their combinations on GISAID. Live analysis from EpiCoV (as of 3 November 2021) containing the results for each of our MOCI, and counts for VOC/VOI according to GISAID. This is compared with data downloaded from GISAID (as of 1 November 2021) and analysed using our in-house alignment pipeline (complete human-origin genomes) after filtering for correct dates in the metadata (‘YYYY-MM-DD’).

|  | **GISAID live analysis using EpiCoV** | **Data downloaded from GISAID (correct dates)** |
| --- | --- | --- |
| **D614G** | 4,245,283 | 4,129,785 |
| **N501Y** | 1,255,861 | 1,220,340 |
| **P681R** | 1,887,719 | 1,844,450 |
| **D614G + N501Y** | 1,251,360 | 1,216,080 |
| **D614G + P681R** | 1,882,323 | 1,839,455 |
| **N501Y + P681R** | 3,728 | 3,688 |
| **D614G + N501Y + P681R** | 3,718 | 3,678 |
| **D614G + N501Y + P681H** | 1,112,530 | 1,082,482 |
| **Alpha** | 1,127,758 | 1,096,665 |
| **Beta** | 39,478 | 37,935 |
| **Gamma** | 96,217 | 92,164 |
| **Delta** | 1,871,122 | 1,828,769 |
| **Alpha + P681R** | 2,393 | 2,352<sup>a</sup> |
| **Beta + P681R** | 24 | 24<sup>b</sup> |
| **Gamma + P681R** | 294 | 294<sup>c</sup> |
| **Delta + N501Y** | 917 | 912<sup>d</sup> |
| <sup>a</sup> mostly present in the sub-lineage Q.4 (2119 samples, since 13 December 2020), B.1.1.7 (230 samples, 17 October 2020) |  |  |
| <sup>b</sup> mostly present in B.1.351 (23 samples, since 30 March 2021) |  |  |
| <sup>c</sup> mostly present in sub-lineage P.1.8 (180 samples, since 20 August 2021) |  |  |
| <sup>d</sup> mostly present in B.1.617.2 (476 samples, since 29 March 2021), sub-lineage AY.33 (155 samples, since 30 April 2021) |  |  |

**Supplementary Table S3.** Observation of other key SARS-CoV-2 Spike mutations. Live analysis from EpiCoV (as of 3 November 2021) containing the results for other key Spike mutations according to GISAID. This is compared with data downloaded from GISAID (as of 1 November 2021) and analysed using our in-house alignment pipeline after filtering for correct dates in the metadata (‘YYYY-MM-DD’).

|  | **GISAID live analysis**<br>**using EpiCoV** | **Data downloaded**<br>**from GISAID** | **Data downloaded from**<br>**GISAID (correct dates)** |
| --- | --- | --- | --- |
| **Total** | 4,835,087 | 4,766,139 |  |
| **Human** | 4,831,865 | 4,763,023 |  |
| **Human complete** | 4,748,462 | *4,294,469 | 4,177,098 |
| **L18F** | 212,488 | 197,955 | 191,238 |
| **T20N** | 108,445 | 93,507 | 89,541 |
| **H69del** | 1,123,439 | 1,138,302 | 1,108,661 |
| **Y145del** | 1,364 | 2,617 | 2,563 |
| **A222V** | 403,457 | 367,047 | 352,024 |
| **K417N** | 44,282 | 43,114 | 41,449 |
| **N439K** | 35,253 | 35,135 | 34,365 |
| **L452R** | 2,304,322 | 1,919,808 | 1,877,945 |
| **Y453F** | 1,255 | 1,242 | 1,221 |
| **G476S** | 933 | 799 | 764 |
| **S477N** | 70,642 | 69,479 | 67,649 |
| **T478I** | 902 | 892 | 879 |
| **V483A** | 296 | 264 | 243 |
| **E484K** | 225,663 | 205,807 | 197,301 |
| **E484Q** | 11,834 | 10,746 | 9,998 |
| **E780Q** | 5,931 | 5,733 | 5,627 |
| **V1176F** | 125,546 | 108,949 | 103,509 |
| *complete human-origin genomes |  |  |  |

**Supplementary Figure S4.**
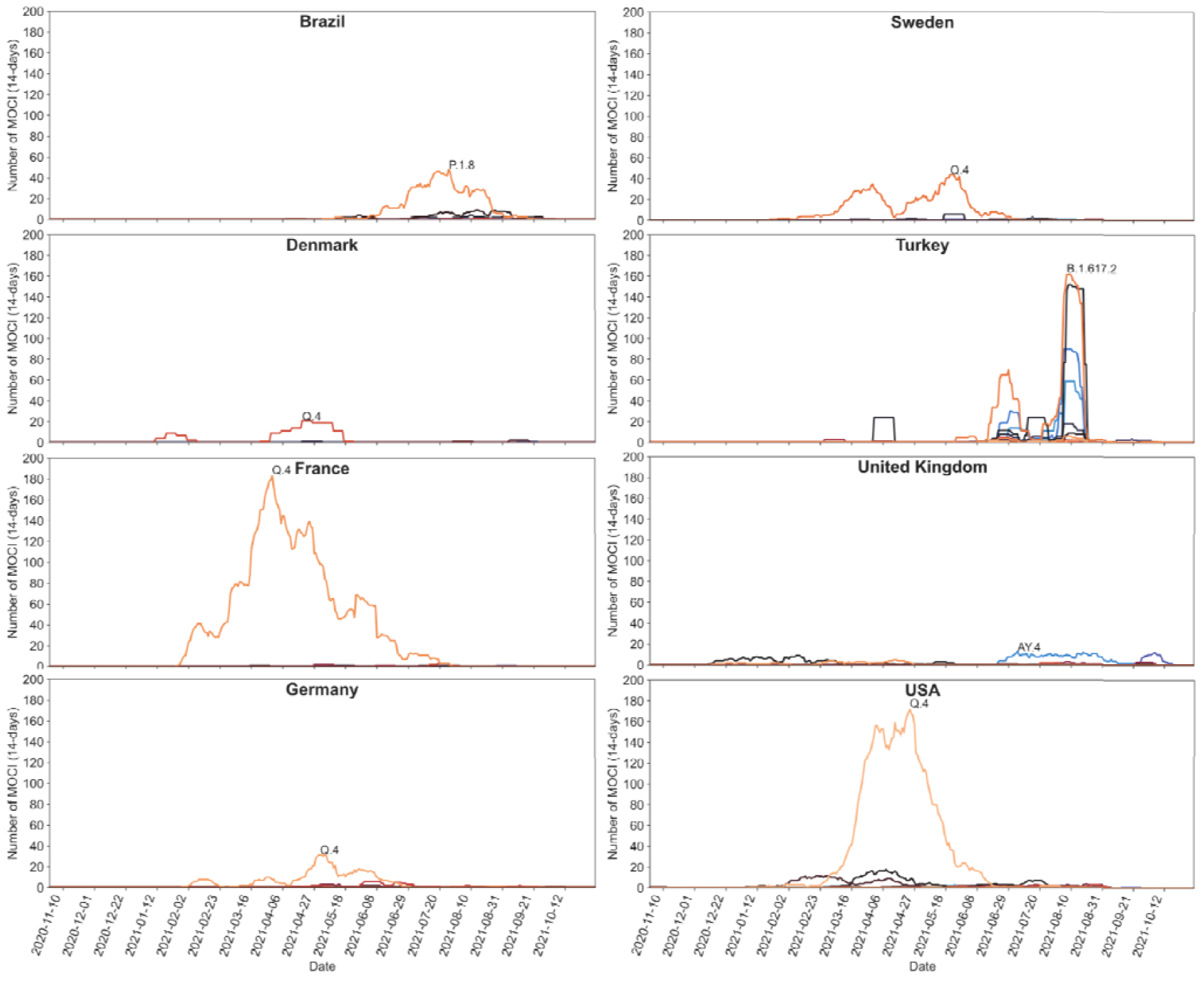
Frequency of the P681R, N501Y and D614G mutations cooccurring in the eight countries with more than 50 observations between October 2020 and October 2021. For visual clarity, only the SARS-CoV-2 lineage with the maximum count is labelled.

**Supplementary Table S5.** GISAID data on key SARS-CoV-2 mutations classified by VOC/VOI. Live analysis from EpiCoV (as of 3 November 2021) containing the results for each of our MOCI, and counts for VOC/VOI according to GISAID. This is compared with data downloaded from GISAID (as of 1 November 2021) and analysed using our in-house alignment pipeline after filtering for correct dates in the metadata (‘YYYY-MM-DD’).

|  | **GISAID live analysis**<br>**using EpiCoV** | **Data downloaded**<br>**from GISAID** | **Data downloaded from**<br>**GISAID (correct dates)** |
| --- | --- | --- | --- |
| **Total** | 4,835,087 | 4,766,139 |  |
| **Human origin** | 4,831,865 | 4,763,023 |  |
| **Human and complete** | 4,748,462 | *4,294,469 | 4,177,098 |
| **D614G** | 4,692,072 | 4,245,283 | 4,129,785 |
| **N501Y** | 1,282,756 | 1,255,861 | 1,220,340 |
| **P681R** | 2,288,985 | 1,887,719 | 1,844,450 |
| **Alpha** | 1,135,705 | 1,127,758 | 1,096,665 |
| **Beta** | 38,865 | 39,478 | 37,935 |
| **Gamma** | 113,091 | 96,217 | 92,164 |
| **Delta** | 2,267,806 | 1,871,122 | 1,828,769 |
| **Lambda** | 8,771 | 8,149 | 8,084 |
| **Mu** | 13,042 | 12,447 | 11,814 |
| *complete human-origin genomes |  |  |  |

## Graphical Abstract

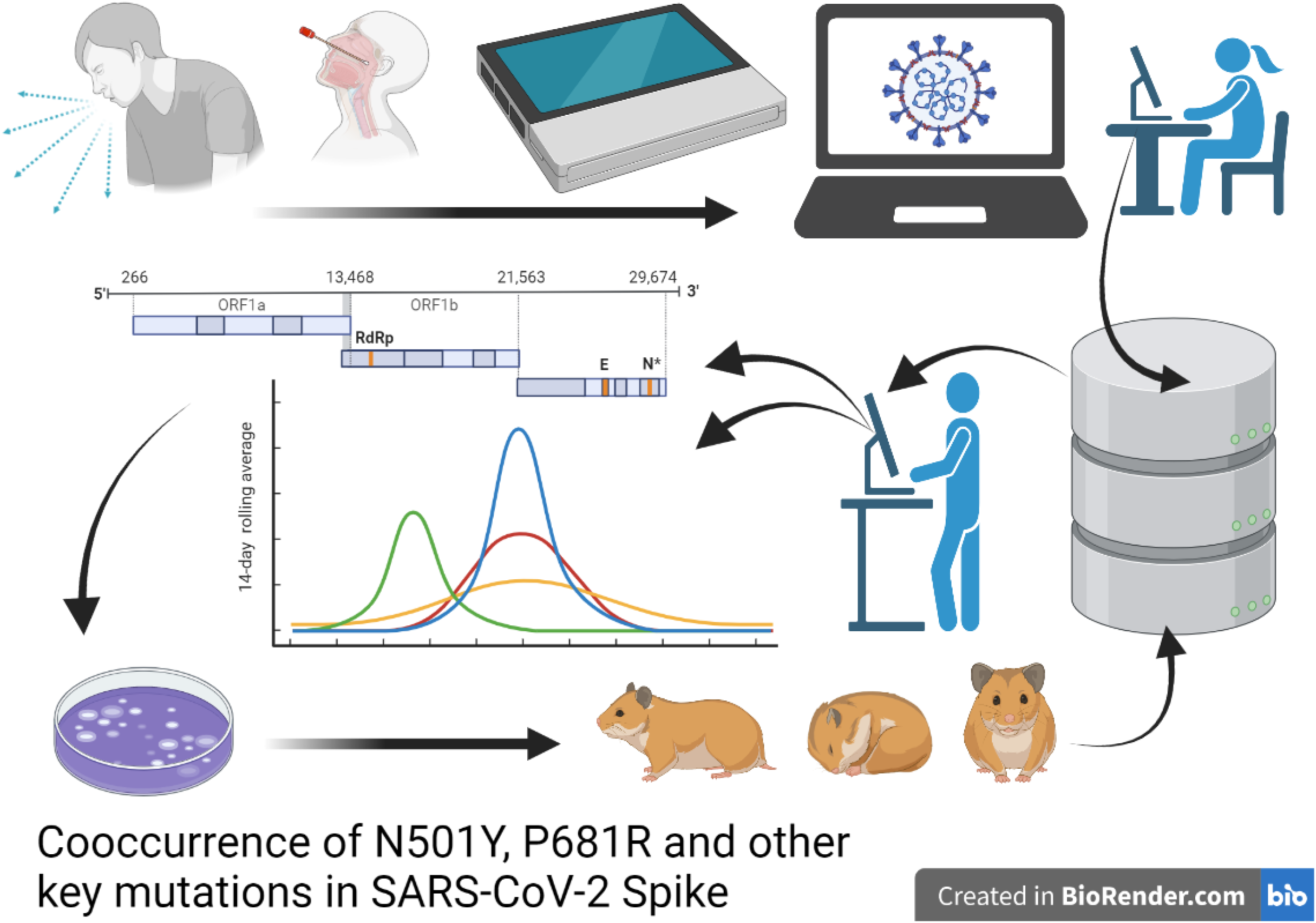

