## Supplementary figures and images for "Cooccurrence of N501Y, P681R and other key mutations in SARS-CoV-2 Spike"

### Supplementary Figure S4

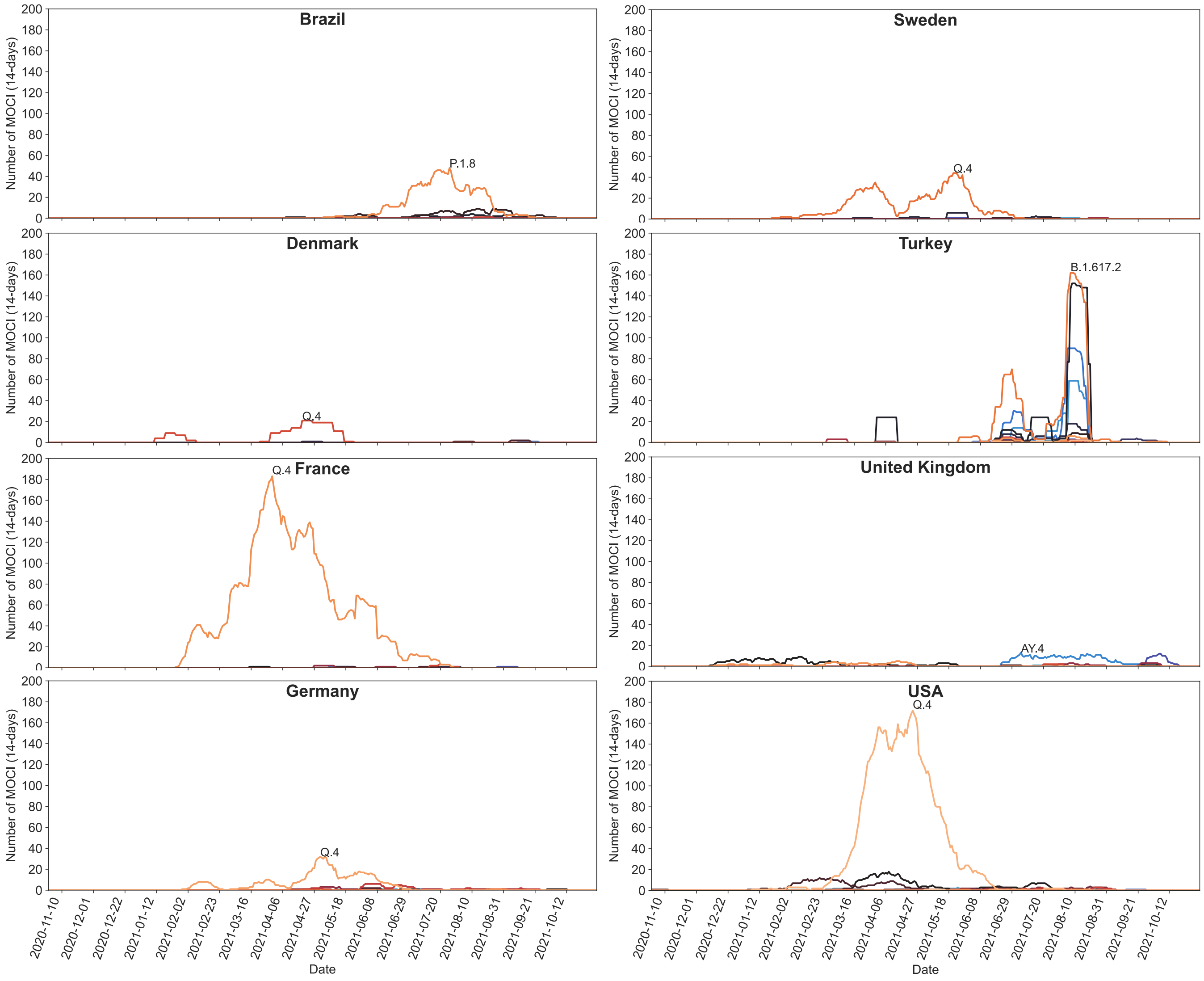
